# Integrated analysis of patient networks and plasmid genomes reveals a regional, multi-species outbreak of carbapenemase-producing Enterobacterales carrying both *bla*_IMP_ and *mcr-9* genes

**DOI:** 10.1101/2021.10.28.21265436

**Authors:** Yu Wan, Ashleigh C. Myall, Adhiratha Boonyasiri, Frances Bolt, Alice Ledda, Siddharth Mookerjee, Andrea Y. Weiße, Maria Getino, Jane F. Turton, Hala Abbas, Ruta Prakapaite, Akshay Sabnis, Alireza Abdolrasoulia, Kenny Malpartida-Cardenas, Luca Miglietta, Hugo Donaldson, Mark Gilchrist, Katie L. Hopkins, Matthew J Ellington, Jonathan A. Otter, Gerald Larrouy-Maumus, Andrew M. Edwards, Jesus Rodriguez-Manzano, Xavier Didelot, Mauricio Barahona, Alison H. Holmes, Elita Jauneikaite, Frances Davies

## Abstract

**Background:** Carbapenemase-producing Enterobacterales (CPE) are challenging in the healthcare setting, with resistance to multiple classes of antibiotics and a high associated mortality. The incidence of CPE is rising globally, despite enhanced awareness and control efforts. This study describes an investigation of the emergence of IMP-encoding CPE amongst diverse Enterobacterales species between 2016 and 2019 in patients across a London regional hospital network.

**Methods:** We carried out a network analysis of patient pathways, using electronic health records, to identify contacts between IMP-encoding CPE positive patients. Genomes of IMP-encoding CPE isolates were analysed and overlayed with patient contacts to imply potential transmission events.

**Results:** Genomic analysis of 84 Enterobacterales isolates revealed diverse species (predominantly *Klebsiella* spp, *Enterobacter* spp, *E. coli*), of which 86% (72/84) harboured an IncHI2 plasmid, which carried both *bla*_IMP_ and the mobile colistin resistance gene *mcr-9* (68/72). Phylogenetic analysis of IncHI2 plasmids identified three lineages which showed significant association with patient contact and movements between four hospital sites and across medical specialities, which had been missed on initial investigations.

**Conclusions:** Combined, our patient network and plasmid analyses demonstrate an interspecies, plasmid-mediated outbreak of *bla*_IMP_CPE, which remained unidentified during standard microbiology and infection control investigations. With DNA sequencing technologies and multi-modal data incorporation, the outbreak investigation approach proposed here provides a framework for real-time identification of key factors causing pathogen spread. Analysing outbreaks at the plasmid level reveals that resistance may be wider spread than suspected, allowing more targeted interventions to stop the transmission of resistance within hospital networks.

**Summary:** This study describes an investigation, using integrated pathway networks and genomics methods, of the emergence of IMP-encoding CPE amongst diverse Enterobacterales species between 2016 and 2019 in patients across a London regional hospital network, which was missed on routine investigations.

## Introduction

Infections by carbapenemase-producing Enterobacterales (CPE) pose a substantial clinical, operational, and financial challenge [1]. These organisms are associated with high morbidity and mortality, and therapeutic options are severely restricted [2]. Carbapenemase genes are frequently carried on plasmids, which can easily transfer between bacterial species [3]. CPE outbreaks involving different bacterial species are often unrecognised, as many plasmids are variable in their gene content and have a broad host range [4]. Outbreaks of Enterobacterales carrying Imipenemase (IMP) gene *bla*_IMP-1_ are mostly sporadic and often localised to specific geographical locations [5, 6]. IMP genes are rarely isolated in the UK, however, the number of IMP encoding Enterobacterales species isolates referred to UK Health Security Agency has been increasing [7].

Colistin and polymyxin B remain the last-line therapeutic agents for CPE in most countries, partly due to lack of access to newer agents; yet colistin resistance is increasing globally. Ten mobile colistin resistance genes (*mcr-1* – *mcr-10*) have been described to date, presenting a substantial global healthcare challenge [8, 9]. Although *mcr* genes are typically associated with phenotypic polymyxin resistance, *mcr-9* does not appear to confer direct colistin resistance [10, 11] and is widespread in a wide range of bacterial species from human, animal and environments [11–14].

Person-to-person contact is a route of transmission for many infectious diseases. Consequently, understanding the patterns of these contacts, especially in healthcare settings, can offer detailed insight for targeted interventions [15]. However, such patient contacts become increasingly complex when incorporating multi-layers of data. Network models provide flexible tool to capture complex interactions (contact patterns) and, offer robust and reproducible methodology that has become widespread across disciplines [16, 17], incorporating both person-to-person transmission through contact networks [18] and spatial spread through networks representing physical locations [19]. So far, few studies utilised network models of patient contacts in combination with detailed bacterial genomic analysis and demonstrated such approach advantages through increasing the detail in outbreak characterisation [20, 21].

Here, we combine plasmid phylogenomic analysis with patient-contact networks to discover the spread of *bla*_IMP_ and *mcr-9* genes among bacterial species and patients in a large hospital network in London, UK over three years, providing valuable insights for the management of CPE in hospital settings.

## Materials and Methods

### Clinical setting

This study was carried out using data from a regional network of London hospitals, comprising seven hospital sites with a total of 2000 inpatient beds, with managerial responsibility assigned to two National Health Service Trusts, and frequent transfers between Trusts and sites for specialist care. Cases were identified from one of these trusts (comprising five hospitals), with microbiology and pathway data for those cases was identified through a shared centralised microbiology laboratory and Electronic Health Records (EHR) system (Cerner, UK). Since June 2015, an enhanced routine CPE screening programme has been implemented in this trust [22]. When a new case of CPE was identified, the patient was isolated in a single room with contact precautions, the bed space and bathroom were terminally enhanced cleaned, and any contacts were re-screened for CPE.

### Isolate collection

CPE isolates were collected from cases identified through rectal screens or clinical sampling between June 2016 and November 2019. Bacterial species were determined using Biotyper MALDI-TOF mass spectrometry (Bruker Daltonics, Germany). One isolate per species was collected from each patient. Susceptibility to 21 antimicrobials was tested using EUCAST disc-diffusion method, and colistin MICs were retrospectively determined using MICRONAUT broth microdilution (BioConnections, UK) for all viable CPE isolates carrying *bla*_IMP_ genes (hereafter, *bla*_IMP_CPE) [23]. Further phenotypic and molecular characterisation of CPE isolates were performed as described in Supplementary Methods.

### Whole-genome sequencing (WGS)

Isolates of *bla*_IMP_CPE were grown aerobically on Columbia Blood Agar (Oxoid Ltd, UK) at 37°C. Genomic DNA was extracted from overnight cultures using GenElute Bacterial Genomic DNA Kits (Sigma-Aldrich, USA). Multiplexed DNA libraries were generated with Nextera XT (Illumina, USA) and sequenced under a 150-bp paired-end layout for a minimum of 100-fold coverage on Illumina HiSeq 4000 systems (Illumina, USA).

### Phylogenomic analysis

Quality control of sequencing reads, *de novo* genome assembly, and genetic characterisation of isolates are described in Supplementary Methods. A neighbouring-joining tree of CPE genomes was generated from pairwise average nucleotide distances using FastANI v1.33 [24]. Plasmid sequences were reconstructed from genome assemblies using MOB-suite v3.1.0 [25]. Reconstructed sequences of IncHI2 plasmids were aligned against IncHI2 plasmid pKA_P10 (GenBank accession: CP044215.1) using Snippy (github.com/tseemann/snippy) to identify genetic variation. A recombination-corrected maximum-likelihood (ML) tree of IncHI2 plasmids was reconstructed from the sequence alignment using IQ-Tree v2.0.3 [26] as implemented in Gubbins v3.2.1 [27]. The date of the most-recent common ancestor of IncHI2 plasmids was estimated using Bactdating v.1.1.1 [28].

### Network analysis

To reveal potential transmission structure, a patient contact network was reconstructed from patients’ movement history (ward locations and time) extracted from EHR data of *bla*_IMP_CPE cases. A contact was defined as an event when two patients were present on the same ward on the same day. Time-aggregated patient contacts were subsequently clustered to reveal groups of patients linked together using the Walktrap community detection algorithm [29]. Contacts were weighted by the time spent together, and a temporal analysis of patient interactions was performed to assess patient roles and positions in transmission. A spatial network of ward/hospital distributions was generated, allowing calculation of in-hospital infectious periods — days spent on the ward prior to implementation of infection prevention and control (IPC) measures, a network structure to determine ward/hospital spread, and a list of highly visited wards according to plasmid genetic clusters.

To investigate if the identified lineages of IncHI2 plasmids represented the transmission of *bla*_IMP_CPE, a Kendall’s rank correlation coefficient was calculated from pairwise phylogenetic distances between IncHI2 plasmids (extracted from the plasmid ML tree) and shortest-path distances between patients (from whom isolates carrying these plasmids were collected) in the contact network (Supplementary Methods).

### Data availability

Illumina reads and draft genome assemblies of 84 *bla*_IMP_CPE isolates were deposited in European Nucleotide Archive under BioProject PRJEB38818. See Supplementary Table 1 for sample information.

### Ethics

This study was carried out in accordance with ethics reference 21/LO/0170 (279677), protocol 21HH6538 Investigation of epidemiological and pathogenic factors associated with infectious diseases.

## Results

### Incidence of *bla*_IMP_CPE

Following the introduction of the enhanced CPE screening programme, *bla*_IMP_CPE was first observed in two Trusts’ hospitals in June 2016 through routine rectal screening. From November 2016, an increasing number of *bla*_IMP_CPE isolates was identified across Enterobacterales species (Figure 1A). The highest incidence of *bla*_IMP_CPE cases occurred between January and July 2019 (Figure 1B). Altogether, *bla*_IMP_CPE isolates were recovered from screening or clinical samples from 116 patients admitted to these five hospitals by the end of November 2019, when new cases rapidly dropped, and subsequent cases were sporadic and infrequent. No ward or service was identified as a potential focus for cross transmission, and no enhanced IPC measures were taken. Only two clusters of cases (5/116 cases) fitted the conventional outbreak definition that ≥2 cases of the same bacterial species with the same resistance mechanism overlapping in time and space. Pulsed-field gel electrophoresis typing of CPE isolates showed similar profiles, suggesting within-hospital transmission. Furthermore, the daily number of occupied beds revealed a continuous burden of patients colonised with *bla*_IMP_CPE (Figure 1C). This burden was particularly evident for patients colonised by *Enterobacter*, with 424 total bed days in the peak month (March 2019) across the hospital network.

**Figure 1.**
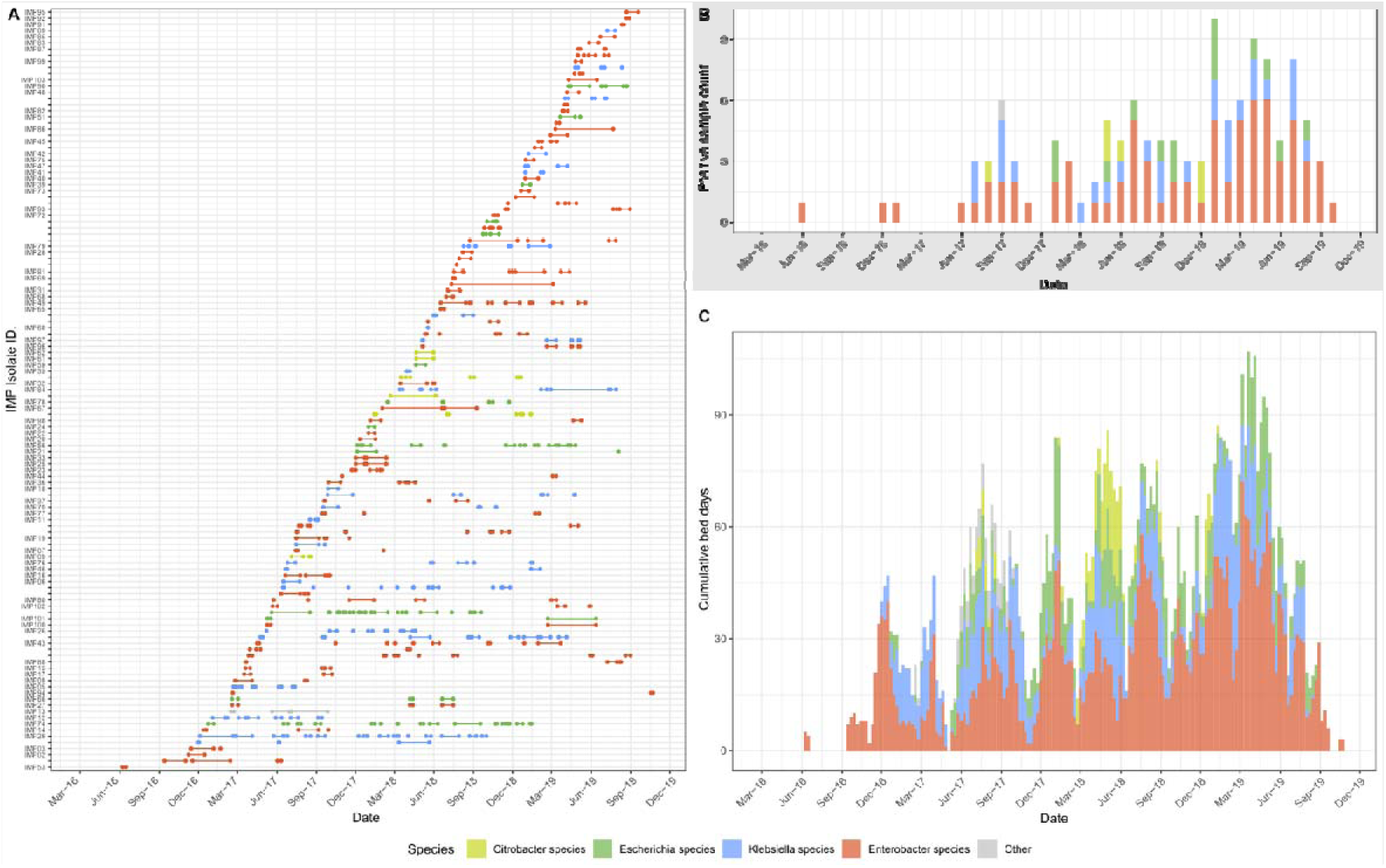
Characteristics of confirmed *bla*_IMP_CPE cases and CPE species. Colours in each panel indicate the genus of CPE. (**A**) Total number of bed days when inpatients (rows labelled by isolate identifier) were present in a hospital ward before confirmation of *bla*_IMP_CPE colonisation/infection and related IPC measures (*in-hospital infectious period*). Additionally, patients with known carriage of *bla*_IMP_CPE but without sequenced isolates are shown as unlabelled rows. Patients with two species of CPE isolates (IMP22/24, IMP25/33, IMP96/97, IMP100/101) are on adjacent rows. (**B**) Monthly total number of confirmed *bla*_IMP_CPE isolates from patients during the study period 2016–2019. (**C**) Weekly cumulative number of occupied beds in-hospital during the infectious periods.

### Contact network of *bla*_IMP_CPE-positive cases

A detailed patient contact network for 116 *bla*_IMP_CPE cases confirmed that 77/116 (66%) cases were in contact with at least one other *bla*_IMP_CPE case (ranged from one to 10 with a median of two cases; Figure 2 and Supplementary Table 2), creating 96 patient-contact pairs (Supplementary Table 3).

**Figure 2.**
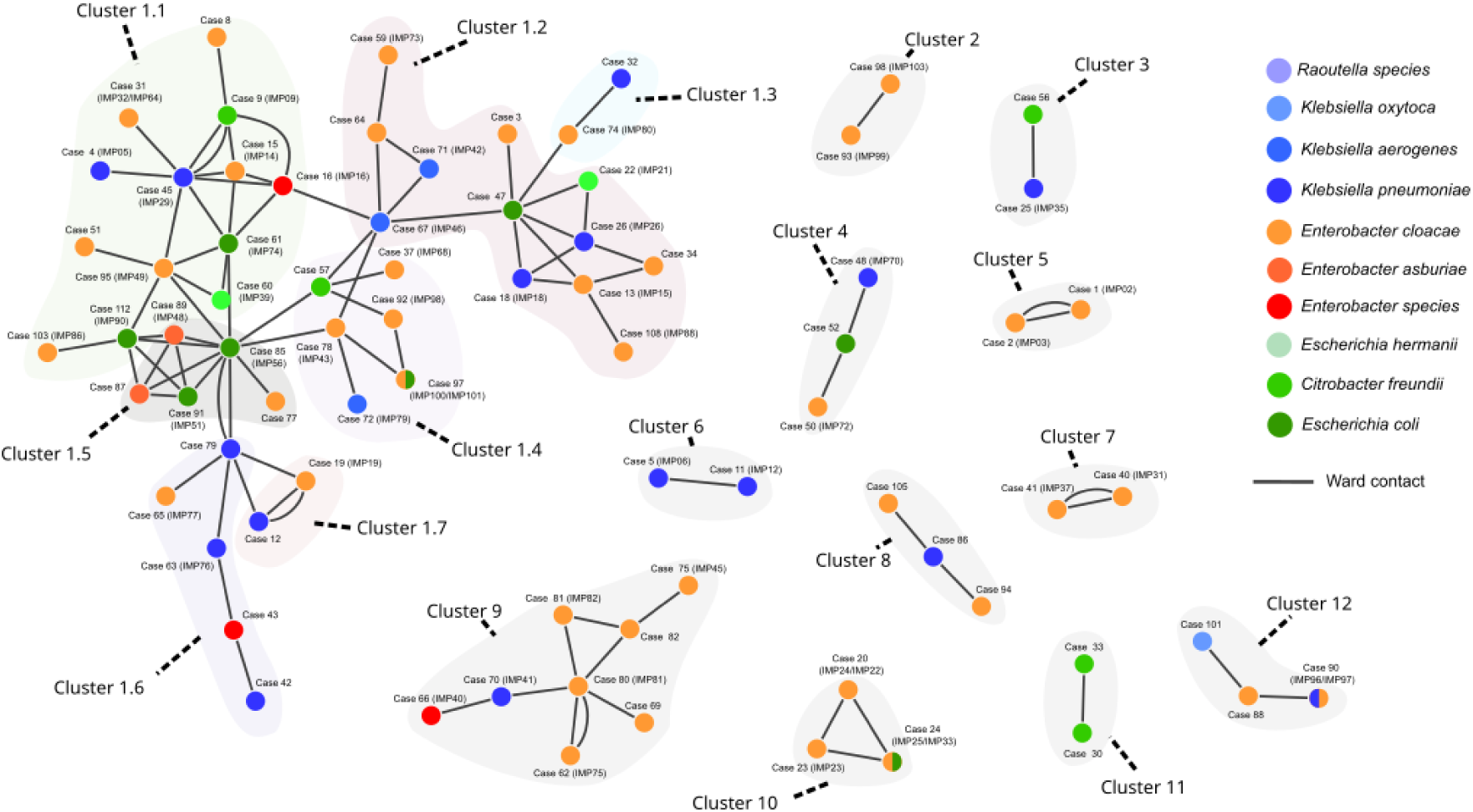
Contact network of *bla*_IMP_CPE cases. Each node of the network represents a case, coloured according to CPE species (split colours indicates two species), and each edge represents a contact between two patients, *i.e.*, patients present on the same ward on the same day based on their electronic health records. This network contains 12 distinct major clusters (each shaded in light grey, with sub-clusters 1.1-1.7 shaded in a different colour) based on disconnected components of contacts. Cluster 1, the largest cluster consisting of 45 cases, was further partitioned into seven subclusters using community detection, with edges weighted by the duration of contact (Supplementary methods – Network community detection). Six patient contacts re-occurred over different wards, indicated by additional edges connecting the same patients.

Across all contact pairs of *bla*_IMP_CPE cases, detected bacterial species differed in 59% (57/96) of patient pairs and therefore, were excluded from the conventional same-species definition of an outbreak when initially reviewed. The network of patient contacts split patients into 12 separate clusters, with interactions occurring across different hospitals, as patients were transferred between wards and hospital sites (Figure 2). The largest contact cluster (Cluster 1) contained 45 patients and was partitioned into a further seven sub-clusters (labelled 1.1 to 1.7) that comprised 13, 12, 2, 6, 5, 5, and 2 patients, respectively (Figure 2). The analysis of contacts at regional, hospital, and ward levels suggested involvement of different *bla*_IMP_CPE species in patient-to-patient transmission events and prompted phylogenomic analysis of available *bla*_IMP_CPE isolates.

### Genomic and phenotypic characterisation of *bla*_IMP_CPE isolates

A total of 84 *bla_IMP_*CPE isolates (collected from 82/116 cases) were available for whole-genome sequencing (Supplementary Table 1). These isolates belonged to 15 species and were dominated by those of the *Enterobacter cloacae* complex (n = 51), followed by *Klebsiella* spp. (n = 21) and *E. coli* (n = 8) (Figure 3). Four cases (Cases 20, 24, 90, 97) were colonised by two *bla*_IMP_CPE species (Supplementary Table 2).

**Figure 3.**
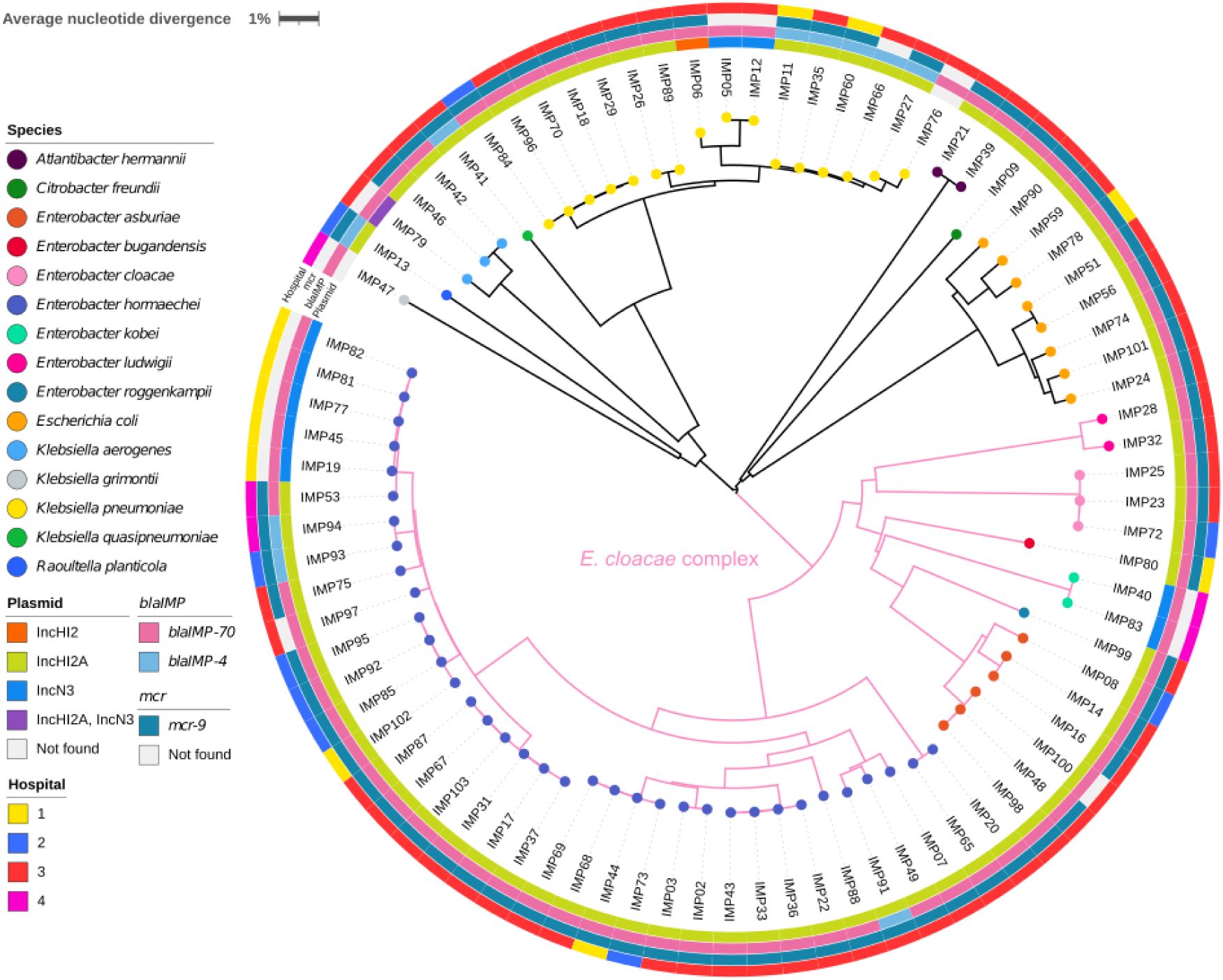
A neighbour-joining tree of 84 *bla*_IMP_CPE isolates. This tree was constructed from average nucleotide distances between genomic sequences and was midpoint rooted. The colour at the end of each branch indicates the bacterial species identity for that isolate. The rings are as follows: the innermost ring indicates the type of plasmid detected; the second ring indicates the allelic variant of the *bla*IMP gene detected, the third ring indicates the presence or absence of gene *mcr-9* and the outermost ring indicates hospitals. The scale bar indicates the pairwise average nucleotide divergence (%).

Each of these 84 isolates carried either *bla*_IMP-70_ (n = 74) and *bla*_IMP-4_ (n = 10). IncHI2 plasmids (targeted by both probes for IncHI2 and IncHI2A replicons in the PlasmidFinder database) were detected in 72 isolates from 72 cases, and IncN3 plasmids were detected in 10 isolates from 10 cases (Figure 3). Only IMP79 harboured both IncHI2 (without any *bla*_IMP_ genes) and IncN3 (carried *bla*_IMP-70_) plasmids. Seventy of the 72 IncHI2 plasmids carried either *bla*_IMP-70_ (n = 61) or *bla*_IMP-4_ (n = 9), and all IncN3 plasmids carried *bla*_IMP-70_ (Supplementary Tables 4 and 5). Four isolates carried *bla*_IMP_ genes that were not found in either IncHI2 or IncN3 plasmids: one *bla*_IMP-4_ was integrated into the chromosome of IMP66, and *bla*_IMP-70_ was carried by an IncFIB/FII plasmid in IMP83 and by IncHI1 plasmids in IMP47 and IMP76. All *bla*_IMP_CPE isolates carried multiple β-lactam resistance genes and other antimicrobial resistance genes, yet only IMP89 had an additional carbapenem-resistance gene *bla*_OXA-48_ (Supplementary Table 1).

Gene *mcr-*9 was detected in 69/84 (82%) isolates, with *mcr-9* identified present on 68 IncHI2 plasmids, one outlier IncHI2 plasmid (32% coverage of the reference plasmid pKA_P10 by sequencing reads), and none of the IncN3 plasmids (Supplementary Tables 4 and 5). The *mcr*-9 LAMP assay showed 100% concordance with the WGS results (Supplemental Table 1). MALDIxin did not detect any Lipid A modifications attributable to the *mcr*-9 gene in this study. Altogether, 12 isolates (all *Enterobacter*) were resistant to colistin (MICs ranged between 4 and >64 μg/mL), including five isolates that demonstrated a skipped-well phenomenon suggestive of colistin heteroresistance (Supplementary Table 1), a phenomenon previously reported [30].

### Genetic relatedness between plasmids

All 72 reconstructed IncHI2 plasmids belonged to the same plasmid taxonomic unit PTU-HI2, and representative sequences are compared in Supplementary Figure 1. Altogether, 144 single-nucleotide polymorphic sites were identified in the alignment of these 72 plasmids after correcting for recombination events, with pairwise phylogenetic distances (sums of branch lengths in the plasmid tree) ranged from zero to 115 single-nucleotide polymorphisms (SNPs). Specifically, of the 72 plasmids analysed in the tree, 43 (60%) differed by ≤3 SNPs, and 55 (76%) differed by ≤5 SNPs. This high degree of similarity between IncHI2 plasmids suggests potential horizontal gene transfer or transfer of full plasmids between different bacterial species. In the case of IncHI2 plasmids present in the same species, a comparison between the plasmid and *E. hormaechei* (the most common species in our data) phylogenetic trees showed likely vertical transmission events as closely related isolates had highly similar plasmids (Supplementary Figure 2). By contrast, reconstructed IncN3 plasmids showed large structural variation in the plasmids (Supplementary Figure 3) and no reliable phylogenetic tree could be reconstructed.

The phylogenetic tree of IncHI2 plasmids indicated three major lineages A, B, and C (Figure 4 and Supplementary Table 6). The estimated date of the most recent common ancestor of the 72 IncHI2 plasmids was 1765 with a large 95% confidence interval of 1536–1895 despite a desirable convergence of the optimised molecular clock model (Supplementary Figure 4), suggesting a lack of temporal signals in reconstructed IncHI2 plasmids.

**Figure 4.**
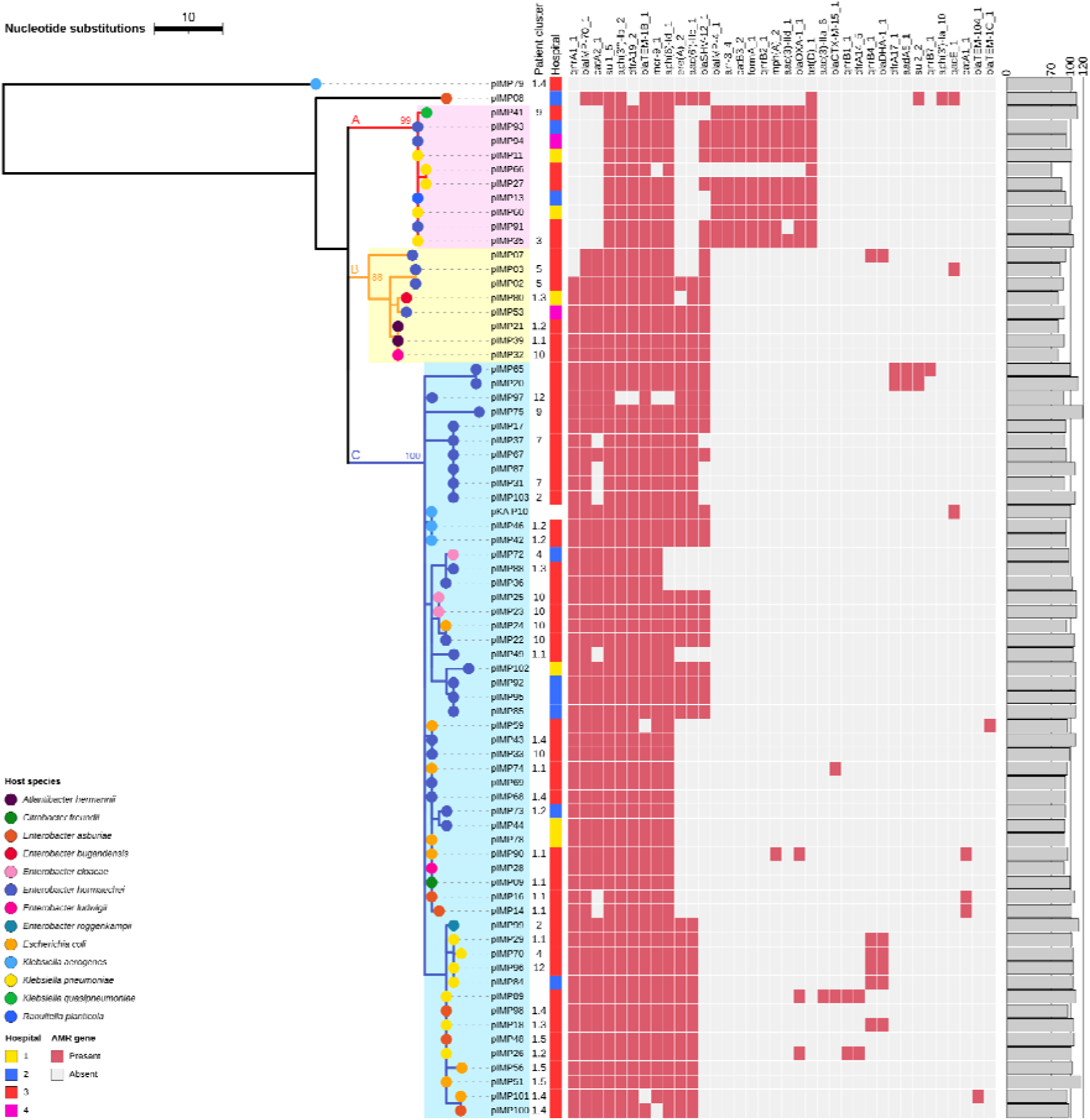
Recombination-corrected maximum-likelihood tree of 72 reconstructed IncHI2 plasmids from *bla*_IMP_CPE isolates and the reference plasmid pKA_P10. Coloured branches and shades represent plasmid lineages A, B, and C, with bootstrap values of lineage roots noted. The heat map shows presence-absence of antimicrobial resistance genes identified in plasmids, and the bar plot shows relative lengths (%) of the reconstructed plasmids compared to that of the reference plasmid pKA_P10. This tree is rooted on the outgroup pIMP79, which was deemed an outgroup according to its phylogenetic distances to other IncHI2 plasmids and by the BactDating root-to-tip analysis (Supplementary Figure 4).

### Comparison between plasmid lineages and patient clusters

Pairwise phylogenetic distances between IncHI2 plasmids and shortest-path distances between patients showed a significant correlation (Kendall’s correlation coefficient = 0.19, p-value = 3×10^−7^) (Figure 5A), despite WGS data being unavailable for isolates from 24 cases. This correlation between plasmid population structure and patient contact network suggests that ward contacts mediated transmission of these plasmids between patients or from unidentified common sources. When case contacts were weighted by patients’ time spent together, the *bla*_IMP_CPE outbreak was heavily weighted towards Hospital 3, the specialist referral centre for cardiology, renal, haematology and hepatobiliary services, with 72.1% of contacts occurring there.

**Figure 5.**
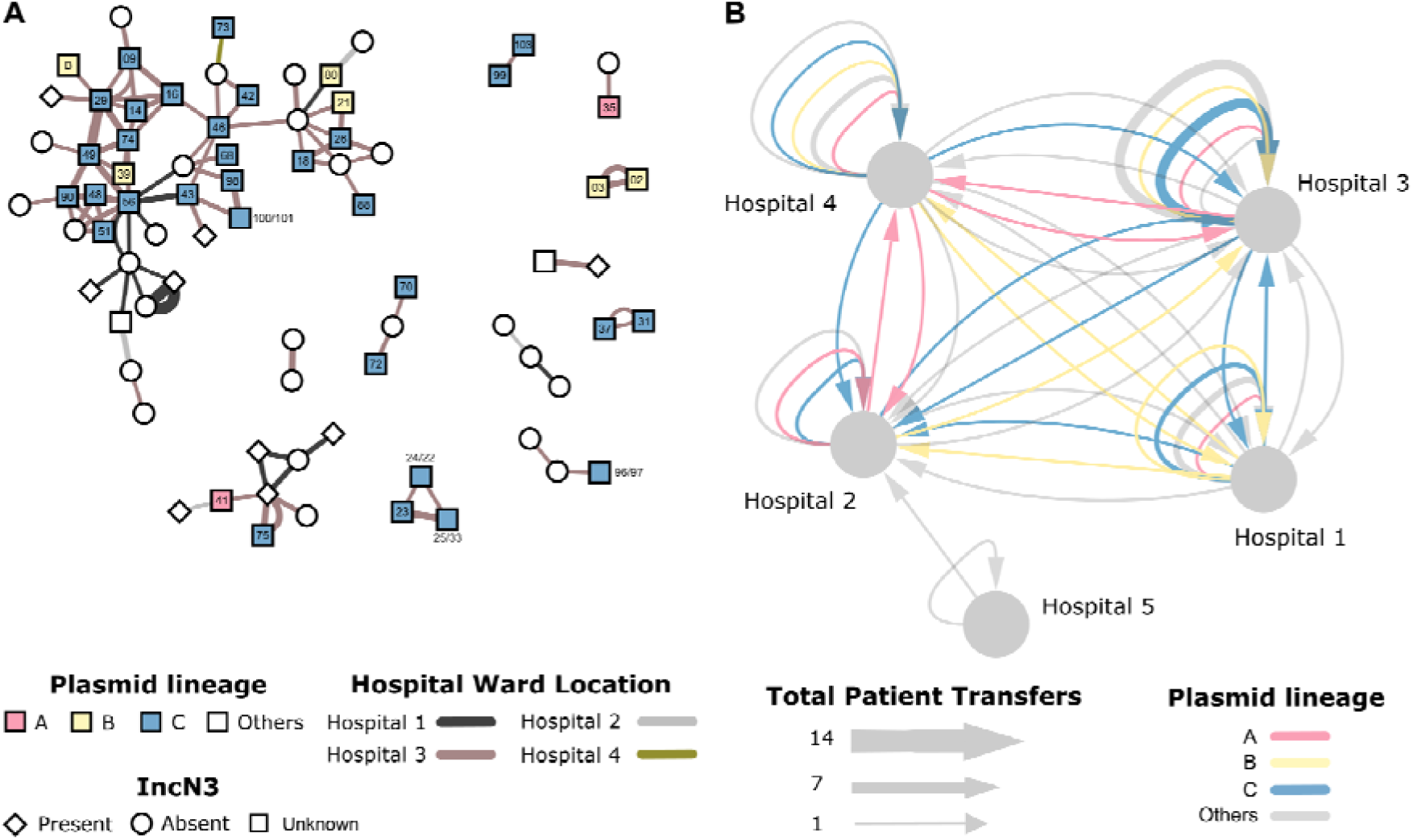
Epidemiology of *bla*_IMP_CPE genetic clusters across patient interactions and movement. (**A**) Patient contact network overlaid with plasmid lineages A, B, and C. Each node represents a patient, edges represent recorded ward contacts between confirmed *bla*_IMP_CPE cases. The edges are coloured according to the hospital site and the width of the edge is proportional to the duration of the contact. Nodes are coloured according to the three lineages of IncHI2 plasmids, and patients with isolates that did not have any IncHI2 plasmids detected are coloured in white. Node labels indicate IncHI2 plasmid names in Supplementary Table 4 (*e.g.*, ‘88’ indicates plasmid pIMP88, and ‘100/101’ indicates plasmids pIMP100 and pIMP101 from the same case). The presence/absence of IncN3 plasmids in *bla*_IMP_CPE isolates is denoted by node shapes. (**B**) Hospital-level patient movements. The movement of patients carrying *bla*_IMP_CPE are indicated by arrows between hospitals. Repeated transfers of patients between wards are aggregated into edges with proportionally greater edge widths (grouped by sequenced and non-sequenced). Edges with sequencing data are coloured according to IncHI2 plasmid lineages.

This was confirmed by the analysis of the spatial distribution and movement of cases colonised with *bla*_IMP_CPE carrying IncHI2 plasmids (Figure 5B). The largest lineage (Lineage C) was found the most prevalent on wards within Hospital 3 (1919 patient bed-days) and followed bidirectional transfer pathways to and from Hospitals 1, 2 and 4, which all have large general medical and surgical admissions areas. Lineage A followed a similar pattern of distribution, though with less transfers identified to Hospital 2, which may have been due to unidentified or missing case data.

The association between plasmid lineage clusters and ward/specialties over the study period showed the most common associations across critical care and renal services (Supplementary Table 7). The only exception was general internal medicine and general surgery predominated in plasmid Lineage A at Hospital 4, which has more general wards and less specialist services than the other hospital sites in the network. Despite the predominance of cases being identified in specialties with high risk for invasive disease, only four clinical infections were identified during the study, and no blood stream infections.

## Discussion

Following the detection of a new mechanism of resistance, investigation of its origin and mode of transmission is challenging, especially in healthcare settings where investigations usually focus on single species transmissions. With confounding factors such as multiple bacterial species and spread over different hospital locations, new methods to investigate potential outbreaks are much needed. The incorporation of plasmid genomics and patient networks into our analysis changed the way the emergence of *bla*_IMP_CPE was visualised and produced a clearer understanding of the cumulative burden of cases, high-risk ward locations and pathways for potential cross transmission in our regional healthcare system. As patients were found to follow common routes, with regular re-encounters, this information can provide dynamic risk assessments to be introduced along those pathways, to prevent future cross transmission events of any healthcare-associated pathogen from occurring [31]. Detailed genomic analysis of plasmids enhanced our understanding of the relatedness of different patient isolates to the network analysis, and similarity to those plasmids identified in other hospitals in the UK [32]. It moreover revealed concerning information about unsuspected resistance mechanisms, with potential for antibiotic treatment failures that were missed on conventional laboratory susceptibility testing. In this study, we characterized IncHI2 plasmids as the main vehicle in horizontal transfer of the metallo-β-lactamase gene *bla*_IMP_. IncHI2 plasmids are common, large plasmids with a wide host range, that have been reported globally [14]. Although in our study we detected *mcr-9* in 81% of the isolates tested, we did not find evidence of phenotypic expression of this gene, in line with previous observations [11, 33]. We identified the predominant IncHI2 plasmid in multiple different bacterial species across linked patients, highlighting the need for integration of genomics into routine clinical practice. Although this study focussed on the emergence of the *bla*_IMP_ carbapenemase gene in our hospital network, it supports the concept that plasmid analysis across different resistance mechanisms as well as among different species should be the standard for investigations in the future. Network analyses and cumulative burden analyses can help identify targets for WGS, particularly where resources are not sufficient to support WGS of all new CPE cases identified. The small number of clinical infections from this outbreak in comparison to other CPE outbreaks from our hospital network [34] and other reports of *bla*_IMP_ CPE [11, 14] is noteworthy, and poses questions about the wider importance of this plasmid and the resistance mechanisms revealed in this study. This observation reinforces the argument that screening for silent carriage of CPE in hospitals is key to preventing spread [35–37], and cautious antimicrobial stewardship is essential to prevent expression of hidden resistance mechanisms [38].

We acknowledge some limitations of our study. Firstly, we did not have long read sequences for plasmids analysed as part of this study. As a result, our plasmid tree may omit some similarities and differences between identified IncHI2 plasmids. Secondly, full pathway data across the hospital during the three years of the outbreak was only available for identified positive cases, not for all patients in the hospitals during the study period. It was therefore not possible to fully establish potential missed cases flagging as close contacts but with potential for missed screening or false negative results. Full pathway movement data for all positive cases identified within our hospital network was available, yet neither pathway details nor genomic data were available for other *bla*_IMP_CPE-positive cases identified in the two other regional hospitals who did not visit our institution, thus reducing the understanding in our analysis. Interactions at other potential hospital locations such as interventional imaging or endoscopy were not examined in this study, nor was environmental sampling performed, which could inform future studies on modes of transmission.

Nevertheless, this study highlights a previously unidentified extent of transmission and thus provides valuable new insights into the spread of an emerging resistance mechanism. Moreover, our novel multi-layered methodology, incorporating plasmid phylogeny with contact network analysis, provides invaluable tools for outbreak investigation that can be generalised to a wide range of scenarios.

## Acknowledgements

We thank the staff of the diagnostic microbiology laboratory of North West London Pathology, for isolate collection and storage. We would also like to acknowledge the support of the Imperial College Healthcare Trust NIHR Biomedical Research Centre (BRC). The Imperial BRC Genomics Facility has provided resources and support that have contributed to the research results reported within this paper. The Imperial BRC Genomics Facility is supported by NIHR funding to the Imperial Biomedical Research Centre. This publication made use of the PubMLST website (https://pubmlst.org/) developed by Keith Jolley (Jolley & Maiden 2010, BMC Bioinformatics, 11:595) and sited at the University of Oxford. The development of that website was funded by the Wellcome Trust.

## Author contributions

Conceptualization: FD, EJ, AH, MB, FB, XD, ME. Data curation: YW, BA, EJ, AM, FD, SM, AW, ME. Formal analysis: AB, AM, YW, MG, FB, FD, EJ, AW, MB, AH, ME. Funding acquisition: AB, FD, AH, ME. Investigation: AB, YW, AM, HA, RP, AL, AA, JT, LM, KM-C, AS, GL-M, JR-M. Methodology: FD, AM, AW, SM, YW, EJ, BA, XD, MB, JO, KH, ME, JT, MG. Project administration: FB, AH, FD, EJ. Resources: FD, AH, AE, HD, KH, JT. Supervision: FD, EJ, AH, MB. Validation: KH, JT, ME, HD, AW, SM. Visualization: AB, AM, YW, EJ, FD. Writing – original draft and further drafts: AB, AM, YW, EJ, FD, FB. Writing – review & editing: All authors.

